# Fast Decisions, Clear Guidance: A Qualitative Study of Trauma Aid Design

**DOI:** 10.1101/2025.08.14.25333716

**Authors:** Lindsay Fountain, Kayla Corredera-Wells, Nicholas Cozzi, Jeffrey M. Goodloe, Jenny M. Guido, Alyssa B. Johnson, Christopher S. Kang, Thomas McNally, Andrea Nevedal, James E. Winslow, Gabriela Zavala Wong, Lacey N. LaGrone

## Abstract

**Objectives:** Although trauma clinical guidance has the potential to improve patient outcomes significantly, they are applied inconsistently across the United States today. Work is needed to improve guidance application among those who treat traumatic injury, including emergency medicine (EM) clinicians. A priority step of such improvement is the visual design of clinical directives, as it can markedly influence EM clinicians’ satisfaction and subsequent usage of guidance. This qualitative study aims to determine EM clinicians’ needs and preferences for the visual design of trauma clinical guidance.

**Methods:** As part of a larger exploratory sequential mixed methods study exploring trauma clinical guidance barriers and facilitators, we conducted semi-structured video conference interviews during August and September 2024 with EM clinicians. Participants were selected through purposeful criteria and snowball sampling to ensure diverse representation across professional and personal characteristics. Interviews were audio recorded and analyzed through rapid qualitative content analysis.

**Results:** Twelve EM clinicians completed interviews, providing feedback on their preferences for the visual design of trauma clinical guidance. Clinicians emphasized the need for simple guidance that can be easily used in a time-pressured setting. Dynamic features such as expandable or “clickable” content can provide deeper explanations of guidance concepts without compromising usability. Flowsheets and visual aids can clarify guidance recommendations. Clinicians asked that recommendations be specific and action-oriented.

**Conclusion:** When designing trauma clinical guidance, authors should promote visual design characteristics that prioritize simplicity, dynamic features, visual aids, and specificity.

**KEY MESSAGES:** *What is already known on this topic:* Trauma clinical guidance has limited usage today amongst EM clinicians for myriad reasons, among them cumbersome visual design practices that do not meet clinicians’ needs or preferences for guidance

*What this study adds:* EM clinicians’ preferences for trauma clinical guidance visual design include prioritizing simplicity, leveraging dynamic features to add depth, using visual cues to enhance navigability, and building recommendations with specificity and actionable steps in mind

*How this study might affect research, practice or policy:* Guidance authors and adaptors should prioritize these attributes as they produce future trauma clinical guidance, further reducing barriers to guidance usage amongst EM clinicians.

## BACKGROUND

Contemporary clinical guidance improves patient outcomes, lowering morbidity, mortality, and patient harm while improving care quality.^1, 2, 3^ However, clinical guidance can be heterogenous and inconsistently applied throughout the spectrum of emergency departments (EDs) in the United States, with guidance adherence ranging from 40-60%.^4, 5^ Emergency medicine (EM) clinicians face multifaceted barriers to using non-EM specialty clinical guidance, from difficulties with finding and accessing guidance to cumbersome-to-use materials. EM clinicians’ frustrations with clinical guidance extend to trauma care. They are often amongst the first to care for injured patients, particularly outside of Level I trauma centers. US trauma mortality rates rose 91% from 2000 to 2020, far outstripping the population growth rate of 17.8%, creating both urgency to reduce trauma fatalities and opportunity for EM clinicians to utilize guidance towards achieving better outcomes.^6^

Understanding EM clinicians’ needs from the visual design aspects of trauma guidance can enhance satisfaction with and usage of guidance. Among the many attributes that define clinical guidance, clarity of presentation has a high correlation with EM clinicians’ overall satisfaction with clinical guidance, emphasizing the need to provide clinicians with visually satisfactory materials.^7^ Recent research by an academic-affiliated, urban Level I trauma center into EM clinicians’ preferences for guidance design revealed a clear desire for concise guidance with clear visual cues leading users through its process.^8^ However, further research is needed into the preferences of EM clinicians working in different practice settings, as well as preferences specific to guidance concerning care for injured patients. This qualitative, interview-based study aims to determine EM clinicians’ needs and preferences for the visual design of trauma clinical guidance.

## METHODS

### Study Design and Sample

As part of a larger exploratory sequential mixed methods study exploring trauma clinical guidance barriers and facilitators, we conducted semi-structured interviews to understand EM clinicians’ preferences for the visual design aspects of trauma clinical guidance. Purposeful criteria and snowball sampling were used to recruit US-based EM clinicians who were in active practice at the time of the interview. To diversify the sample, we aimed to include EM clinicians across gender, race, ethnicity, geographic region, trauma center level, clinician profession, years in practice, and level of urbanity.^9^ Geographic regions were determined using Centers for Disease Control and Prevention (CDC) definitions.^10^ Clinicians were recruited via email outreach, leveraging the research team’s network of professional contacts and, as the research progressed, referrals from past participants. Although some participants were known to the project team at the time, none had preexisting relationships with the interviewer, and their identities were blinded to the project team as analysis began. Thirty-four individuals were contacted, twenty-two declined, and twelve completed interviews. The convenience sample cohort size of twelve was sufficient, given the high information power yielded by the study’s narrow aims, dense sample specificity, and strong dialogue.^11^ (Supplemental Item 1 provides additional methods description.)

The project research team included diverse perspectives from the trauma and healthcare research communities, including two trauma surgeons (JG, LL), four emergency medicine physicians (CK, JG, JW, NC), one research physician (GZ), one emergency medicine physician assistant (TM), one state trauma system manager (AJ), one medical anthropologist (AN), and two medical students (KC, LF). This study was reviewed by the Colorado Multiple Institutional Review Board, CB F490; COMIRB #: 24-0047 and determined exempt; interviewees provided verbal consent. No compensation was given for participation in interviews.

### Data Collection

The interview guide was pilot tested amongst coauthors and two medical students not associated with the research team. Interviews were held and recorded via video conference during August and September 2024. The average interview duration was 51 minutes, with a range of 31 to 63 minutes. They were conducted by a medical student (LF) who received prior training in qualitative interviewing techniques from a medical anthropologist. Interviewers encouraged participants to provide candid responses and asked clarifying questions to ensure their perspectives were understood and documented.

In addition to open-ended questions about their preferences for guidance, clinicians were asked to comment on two examples of existing trauma clinical guidance (see Supplemental Item 2 for complete interview guide). The two examples were chosen to include protocols routinely used in the care of trauma patients and to provide a variety of visual styles for participant reaction. The protocols shared were the 2023 American Association for the Surgery of Trauma (AAST) / American College of Surgeons Committee on Trauma (ACS-CoT) Protocol for Damage Control Resuscitation of the Trauma Patient (Figure 1) and the University of California San Francisco (UCSF) Adult Massive Transfusion Protocol (Figure 2, version accessed August 12, 2024).^12,13^ To minimize any potential bias against specific authors, the protocols had any indicators of authorship removed before sharing with participants (see versions below). Example protocols were emailed to participants at least 48 hours before their interview with a request to review the materials and prepare to comment on their strengths, weaknesses, and compatibility with the participant’s current practice setting.

**Figure 1.**
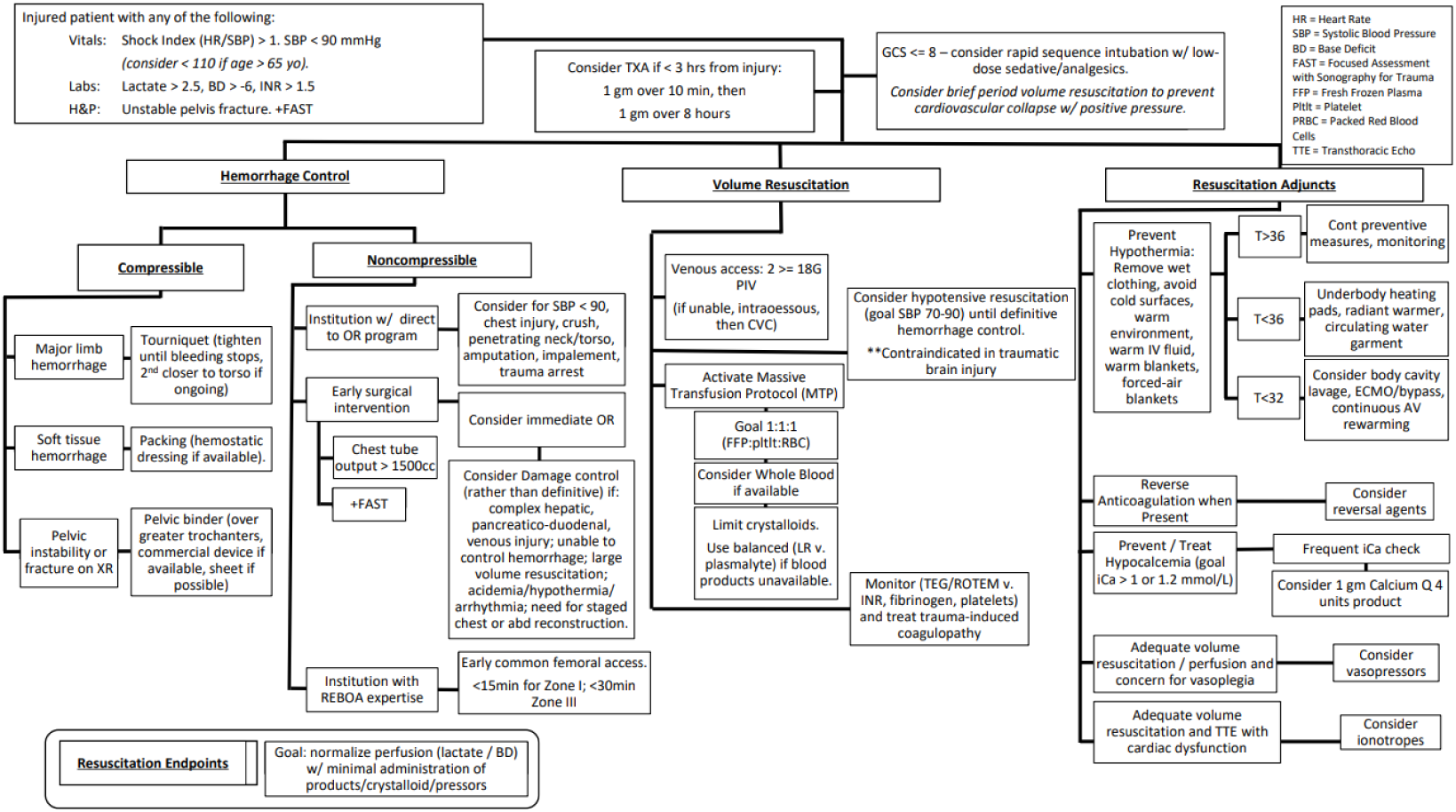
Version of AAST / ACS-CoT Protocol for Damage Control Resuscitation of the Trauma Patient shared with participants.

**Figure 2.**
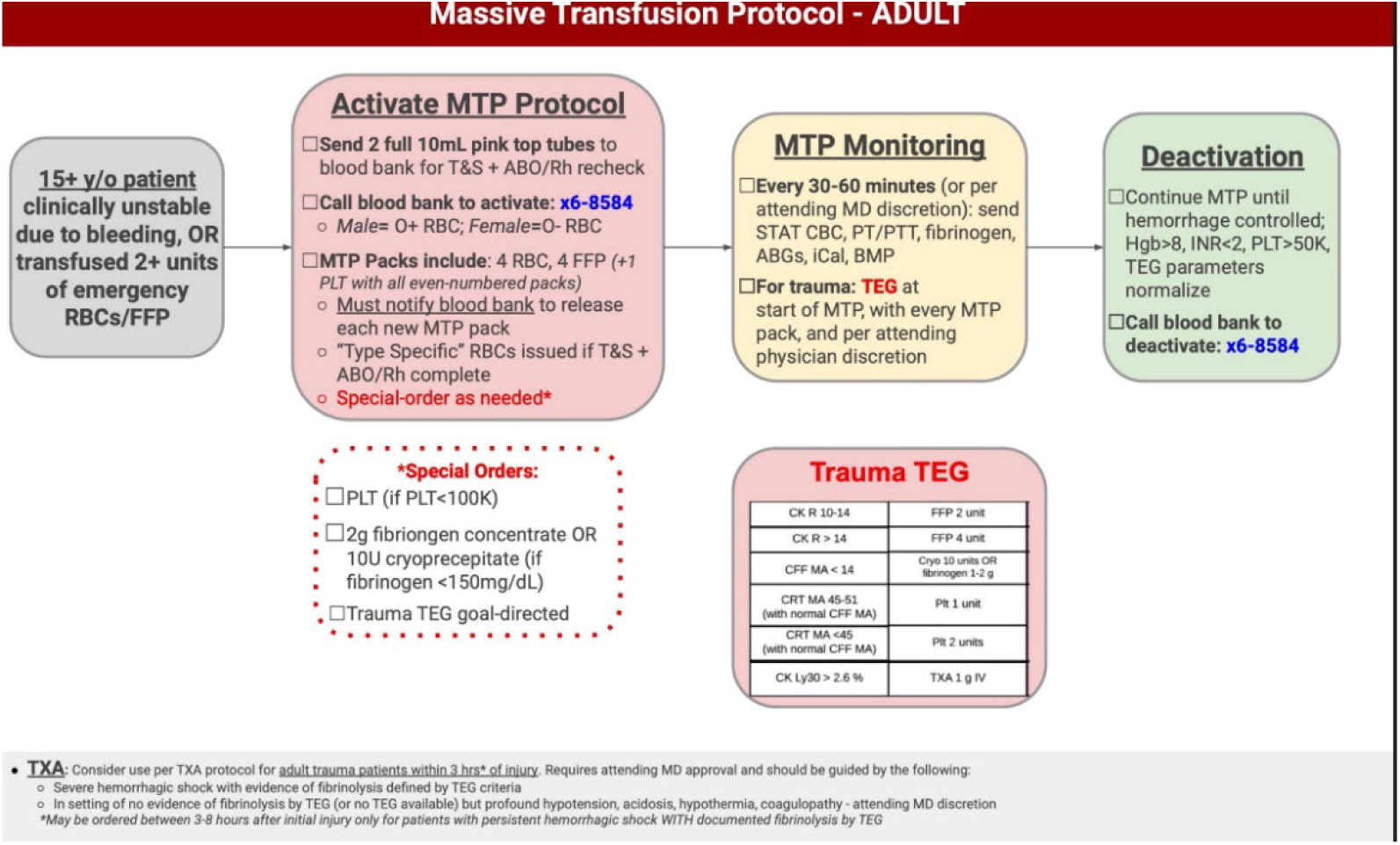
Version of UCSF Adult Massive Transfusion Protocol shared with participants.

### Data Analysis

Interviews were audio recorded and reviewed by two analysts (KW, LF) using rapid directed qualitative content analysis, with methodological expertise provided by a medical anthropologist (AN).^14^ The interviewer and primary analyst (LF) took notes on a template created in Microsoft Office Word™ (Version 2106, Build 14131.20332), with fields to capture responses to each question. The primary analyst then used these notes to draft an interview summary immediately after each interview. The secondary analyst (KW) listened to the audio recordings of each interview to verify the accuracy of and edit each interview summary. The primary analyst then copied the notes into a Microsoft Office Excel™ (Version 2106, Build 14131.20332) matrix containing rows for each participant and columns for domains for each interview question. A Microsoft Word domain summary template was populated for each domain by reviewing the full matrix for relevant quotes. Each domain was analyzed and summarized by comparing and contrasting participant responses for salience, repetition, and negative cases to understand key suggestions for improving trauma visual design.

## RESULTS

Twelve clinicians completed semi-structured interviews (Table 1). Two clinicians regularly practiced in two different settings and shared insights on their perspectives from both locations. The following sections describe the wide range of feedback about guidance shared by clinicians, which is summarized into four suggestions for improvement in guidance design.

**Table 1.** Participant demographics.

| Measure | Item | Count | Percentage (%) |
| --- | --- | --- | --- |
| Clinician profession | Physician | 10 | 83 |
|  | PA | 2 | 17 |
| Clinical duties | Bedside | 9 | 75 |
|  | Administrator | 3 | 25 |
| Years of experience | <10 years | 4 | 33 |
|  | 10-20 years | 3 | 25 |
|  | 20+ years | 5 | 42 |
| Trauma center level | Level I | 4 | 33 |
|  | Level II | 3 | 25 |
|  | Level III | 4 | 33 |
|  | Level IV | 2 | 16 |
|  | Level V | 1 | 8 |
| Degree of urbanity | Urban | 6 | 50 |
|  | Suburban | 4 | 33 |
|  | Rural | 3 | 25 |
|  | Frontier | 1 | 8 |
| Geographic region | Northeast | 2 | 16 |
|  | South | 2 | 16 |
|  | Midwest | 1 | 8 |
|  | West | 7 | 50 |
| Gender | Female | 6 | 50 |
|  | Male | 6 | 50 |
| Race and/or ethnicity | Asian | 1 | 8 |
|  | Black | 3 | 25 |
|  | Hispanic | 2 | 16 |
|  | White | 6 | 50 |

**Table 2.** Summary of emergency medicine clinician needs from trauma guidance visual design.

| Guidance Visual Design Need | Example/explanation |
| --- | --- |
| Design clinical guidance for quick reference and simplicity | Long, wordy guidance is not usable in the time constraints of the ED<br>• "Nothing is gonna stop for you to read a 20-page document. Nothing in the ED is going to stop, particularly when you have these traumas. Time is of the essence. Time is against you." - P12<br>• "It's a lot of information...That's great, but in a crunch situation, will I have to read all of this?" - P7<br>Keep recommendations as succinct as possible by trimming extraneous wording and/or explanations, as well as utilizing de-cluttering formatting (e.g., bullet points)<br>• "Sentences are hard to read. ER people, we think in bullets." - P6 |
| Use dynamic features to support in-depth clinical reasoning | Dynamic features can add detail to clinical guidance without compromising fast usage<br>• "[Guidance should be] flexible and dynamic, with tables and panels that can expand with more information. But it needs to stay on the same page. Once you have multiple windows open and have to go back, it can get messy. It should be like an accordion that opens and closes." - P10<br>• "I think that if you're trying to give somebody a guideline that's quick and dirty and can be used, I think that you can provide that [references and explanations] but it would have to be like, put a star and then it's got to be on the backside or the second page, or something. Explanations can be included, but it should be off to the side." - P9 |
| Leverage flowsheets and visual aids to clarify clinical recommendations | Flowsheets are an effective way to organize trauma clinical guidance recommendations |
|  | • [Flowsheets are helpful because] “trauma when done right is an algorithm” - P5<br>Use arrows, color and intentional sequencing of boxes and branchpoints to guide users through the guidance<br>• “[reacting to Figure 1] I wish it had colors because the flow breaks down a lot. So I think color-coding would help with that. But we shouldn’t use red because some people are colorblind.” - P12<br>• “I like the color. The color helps differentiate where you are in treatment. With the red, that shows where you need the most resuscitation. Green is once things are calm and stable. It tells you okay, now we can move on to the next color.” - P9 |
| Build guidance with specific, action-focused recommendations | Provide objective criteria for decision-making (e.g., not “soft hemodynamics” or “unstable pelvis”)<br>• “[referring to Figure 1] There are some subjective things like “unstable pelvis”. That is difficult to interpret compared to a GCS < 8. An objective equivalent would probably be specific radiology findings that are associated with an unstable pelvis.” - P3<br>Distill guidance to its core components: patient population inclusion criteria, interventions, monitoring, and end points.<br>• “[referring to Figure 2] It clearly defined a patient population, it defined when you would utilize this, and it gave actual specifics for how to accomplish this... who do you call, what do they need, what to say over the phone, and once you get |
|  | the product in, where do you go from there... I like the very specific endpoint.” - P3<br>Ease translation of recommendations into action with institution-specific implementation details (e.g., blood bank phone number, links to order sets)<br>• “[referring to Figure 2] It includes little things like the number for the blood bank. They usually just say ‘call blood bank’ but no one knows the number for the blood bank.” - P4 |

### Design Clinical Guidance for Quick Reference and Simplicity

Clinicians emphasized concerns about guidance being too time-consuming to read and apply, given the unique time constraints of emergency medicine. An exemplar quote to support this concern was:

> “Nothing is gonna stop for you to read a 20-page document. Nothing in the ED is going to stop, particularly when you have these traumas. Time is of the essence. Time is against you.*“* (P12)

While thoroughness added value to the guidance, it is likely outweighed by the need for speed in the acute care setting (e.g., “It’s a lot of information…That’s great, but in a crunch situation, will I have to read all of this?” - P7). Clinicians also reported that textually dense guidance created a substantial initial hurdle for using guidance. For example, clinicians were “turned off right away” (P1) by guidance that was multiple pages long. Clinicians remarked that if offered “dense, 10-page trauma guidelines” (P8), no one would use them.

When clinicians were asked what should be avoided when creating clinical guidance, nine clinicians mentioned verbally dense guidance. The high numbers of words “make people nervous” (P1) and paragraphs of text seemed to detract from usability. Bullets were the preferred format for delivering any verbal information. Paragraphs and even longer sentences created friction for guidance users (e.g., “Sentences are hard to read. ER people, we think in bullets” - P6). Other participants described redundancy and repetitiveness as characteristics to avoid.

### Use Dynamic Features to Support In-Depth Clinical Reasoning

Despite a general aversion to detailed, longer-form guidance, clinicians acknowledged that the style of guidance presentation can add value when used outside the context of point-of-care. Examples of alternative uses for denser guidance similar to Figure 1 included “looking at it as a process map between shifts, to study and know it later” (P10). The concepts described in the guidance could be applied during a shift, but clinicians would need to be familiar with the guidance before the relevant patient encounter arose.

Preferable to guidance that needed to be reviewed ahead of time was the concept of one-page, streamlined guidance that contained more detail outside of the main “quick and dirty” portion of the guidance (P9). More references and explanations may be helpful for clinicians to review, but they should be off to the side or located on a second page.

Clinicians suggested that incorporating dynamic elements into guidance design could add depth and detail to guidance without cluttering the “need-to-know” information included in a point-of-care-focused piece of guidance. Clinicians desired guidance “to be flexible and dynamic, with tables and panels that can expand with more information. But it needs to stay on the same page. Once you have multiple windows open and have to go back, it can get messy. It should be like an accordion that opens and closes” (P10). Others requested “clickable links where bullets popped up with more resources” (P6) or capabilities in the electronic medical record to hover over parts of the guidance to elaborate on a specific point.

### Leverage Flowsheets and Visual Aids to Clarify Clinical Recommendations

All clinicians described their preferred visual format for guidance to be a flowsheet like the ones featured in Figures 1 and 2. One clinician explained that “trauma when done right is an algorithm” (P5), and the “if-then” nature of boxes and arrows captured that dynamic well. Clinicians reported that flowsheets were common in emergency medicine, and that familiarity allows clinicians to read and interpret guidance more quickly. Infographic-style formatting with diagrams may have a place for clinical procedure guidance, but flowsheets are more amenable for any guidance concerning decision making.

Though clinicians favored flowsheets overall, they cautioned that crowded guidance with limited visual cues regarding workflow still prove difficult to interpret in the acute setting. Flowcharts need to be “as simple as can be” and that “trying to tamp through 60 boxes will make people flustered” (P1). Complex flowcharts such as Figure 1 were found to be “overwhelming and [it is] hard to know where to start” (P7). Another clinician knew where the starting point was but found the large number of branch points made it “hard to keep track of where I’m going next” (P10). Arrows helped clinicians understand the flow of each document, but only when moving in one direction and typically from left-to-right.

Clinicians found many of the style elements featured in Figure 2 increased navigability. Several highlighted the use of color as a quick way to draw the eye in and to delineate each phase of the workflow (e.g., “Green means go, you’re at your endpoint. Red is for the unstable patient. Yellow means in process*.”* - P7).

However, clinicians acknowledged that overreliance on color cues would disadvantage colorblind guidance users. Other, less color-centric suggestions for improving navigability included increasing white space between boxes of text and judicious use of capitalization and bold fonts.

### Build Guidance with Specific, Action-Focused Recommendations

Objectivity in patient inclusion criteria and decision-making points increased guidance value to clinicians, particularly for those with less experience. Examples included replacing subjective criteria such as “unstable pelvis” with an objective criterion such as radiographic findings that indicate pelvic instability or giving clear blood pressure ranges rather than stating “soft hemodynamics” (P3). Though overly strict criteria (e.g., specific GCS scores) constrain clinician decision making, most found that providing ranges or multiple ways for patients to meet criteria strikes a balance between specificity and flexibility.

Distilling guidance to action-oriented components reduced friction for clinicians translating recommendations into practice. Clinicians identified patient population inclusion criteria, interventions, monitoring, and defined endpoints as essential components for complete guidance. Objectivity improved usability for each of these components, from definitions for hemorrhage control to specific metrics for monitoring to deactivation criteria. Hospital-specific details for application of guidance during patient care also eased use for clinicians. Generic recommendations such as “call blood bank” frustrated clinicians seeking to act quickly (e.g., “no one knows the number for the blood bank” - P4), and details such as including the current number for the blood bank on the guidance help speed action in patient care. Other measures, such as linking recommendations to order sets or providing specific dosing for medications, also decrease the time to implementation for interventions.

## DISCUSSION

Our study identifies methods for improving the visual design of trauma clinical guidance so that end users, including EM clinicians, can effectively use guidance to improve patient outcomes. Clinical guidance authors, as well as those adapting guidance to a specific hospital system or practice setting, should incorporate the following as they format their recommendations: 1) design for quick reference, 2) use dynamic features, 3) leverage flowsheets and visual aids, and 4) build specific, action-oriented recommendations. Though little visual design research specific to trauma guidance in the EM setting exists to date, our findings are congruent with past findings from research into overall EM guidance needs^8^. EM clinicians continue to call for clearer visual design and simplicity as means to increase usage in time-pressured practice settings. A newer finding from our work is the growing potential for dynamic, expanding features in guidance to provide optionality for clinicians seeking to deepen their clinical reasoning without sacrificing simplicity during point-of-care use. As electronic resources become more widespread, the capability grows for teams to incorporate these dynamic features into trauma clinical guidance.

This work represents a focused, rapid project within a larger mixed methods study. Future work could include a larger study focusing on a broader range of questions about trauma clinical guidance. Participants were generally balanced across their professional and personal characteristics, but there was a disproportion of participants in the West geographic region relative to the rest of the United States and of physicians relative to other clinician professions. No military-affiliated practice settings were included. Additional interviews with clinicians from other regions, professions, and practice settings could impact our understanding of the guidance needs.

The goal of this qualitative study was to obtain rich insights from EM clinicians about their needs and preferences for visual design. We will build off this work to develop a national survey to validate and further explore trauma clinical guidance needs, including but not limited to visual design. Beyond the survey, other future work could include user experience testing of guidance developed using the visual design recommendations distilled in this work.

## CONCLUSION

In a qualitative study of EM clinicians, we identified four methods for improving the visual design of trauma clinical guidance. Given the importance of visual design in clinician satisfaction with guidance, implementing these changes in future versions of guidance may increase EM clinicians’ usage of and satisfaction with trauma clinical guidance. As many grapple with how to reduce the rising burden of trauma mortality in the United States, improvements in trauma clinical guidance should play an important role in those efforts.

## Data Availability

Limited (and deidentified) data produced through the present study are only available through reasonable request to the authors.

## Author Contributions as per CRediT (https://credit.niso.org/)

Fountain - formal analysis, investigation, methodology, project administration, writing – original draft; Corredera-Wells – data curation, formal analysis; Cozzi - writing - reviewing & editing; Goodloe - writing - reviewing & editing; Guido - writing - reviewing & editing; Johnson - writing - reviewing & editing; Kang - writing - reviewing & editing; McNally – resources, writing - reviewing & editing; Nevedal – methodology, writing – reviewing & editing; Winslow - writing – reviewing & editing; Zavala Wong - writing – reviewing & editing; LaGrone – conceptualization, supervision, writing – reviewing & editing, and guarantor

## Human Subjects Statement

This study was reviewed by the Colorado Multiple Institutional Review Board, CB F490; COMIRB #: 24-0047, and determined exempt from institutional review board.

## SUPPLEMENTAL ITEMS

### Supplemental Item 1. Consolidated criteria for reporting qualitative studies (COREQ)

| No. Item | Guide description | Explanation | Reported on Page # |
| --- | --- | --- | --- |
| **Domain 1: Research team and reflexivity** |  |  |  |
| *Personal characteristics* |  |  |  |
| 1. Interviewer/facilitator | Which authors conducted the interview or focus group? | Lindsay Fountain | 6 |
| 2. Credentials | What were the researchers' credentials? | BS | - |
| 3. Occupation | What was their occupation at the time of the study? | Medical student | 6 |
| 4. Gender | Was the researcher male or female? | Female | - |
| 5. Experience and training | What experience or training did the researcher have? | LF was trained in qualitative interviewing techniques by a medical anthropologist. | 6 |
| *Relationship with participants* |  |  |  |
| 6. Relationship established | Was there a relationship established prior to study commencement? | No | 6 |
| 7. Participant's knowledge of the interviewer | What did the participants know about the researcher? E.g. personal goals, reasons for doing the research | Participants were informed on the background and goals of the study before the interview | - |
| 8. Interviewer characteristics | What characteristics were reported about the interviewer/facilitator? e.g. Bias, assumptions, reasons and interests in the research topic | LF is a medical student with experience working as a scribe in two emergency departments | - |
| **Domain 2: study design** |  |  |  |
| *Theoretical framework* |  |  |  |
| 9. Methodological orientation and theory | What methodological orientation was stated to underpin the study? e.g. grounded theory, discourse analysis, ethnography, phenomenology, content analysis | We conducted rapid qualitative content analysis of interview notes and audio recordings. | 6-7 |
| *Participant selection* |  |  |  |
| 10. Sampling | How were participants selected? | Eligible participants included active EM clinicians working in the United States. Potential participants were initially identified using the coauthors' professional networks. Purposeful criteria and snowball sampling further informed selection. | 6 |
| 11. Method of approach | How were participants approached? e.g. face-to-face, telephone, mail, email | Participants were contacted via email | 6 |
| 12. Sample size | How many participants were in the study? | 12 participants in total | 8 |
| 13. Non-participation | How many people refused to participate or dropped out? Reasons? | 22 participants who received our invitations did not respond or were not able to schedule an interview due to time constraints. | - |
| *Setting* |  |  |  |
| 14. Setting of data collection | Where was the data collected?<br>e.g. home, clinic, workplace | Data was collected via video conference | 6 |
| 15. Presence of non-participants | Was anyone else present besides the participants and researchers? | Interviews were conducted without anyone else present | - |
| 16. Description of sample | What are the important characteristics of the sample? e.g. demographic data, date | Described in Table 1 | 6,9 |
| *Data collection* |  |  |  |
| 17. Interview guide | Were questions, prompts, guides provided by the authors? Was it pilot tested? | The interviews utilized a semi-structured format. Questions are shared in Supplemental Item 2. | - |
| 18. Repeat interviews | Were repeat interviews carried out? If yes, how many? | No | - |
| 19. Audio/visual recording | Did the research use audio or visual recording to collect the data? | All interviews were audio-recorded. | 6 |
| 20. Field notes | Were field notes made during and/or after the interview or focus group? | Yes, field notes were made during data collection and checked for accuracy by a secondary analyst during analysis. | 7 |
| 21. Duration | What was the duration of the interviews or focus group? | The average interview duration was 51 minutes, ranging from 31 to 63 minutes. | 6 |
| 22. Data saturation | Was data saturation discussed? | Information power was prioritized over data saturation. The sample size of twelve was sufficient given the high information power yielded by the study's narrow aims, dense sample specificity, and strong dialogue. | 6 |
| 23. Transcripts returned | Were transcripts returned to participants for comment and/or correction? | No | - |
| **Domain 3: analysis and findings** |  |  |  |
| *Data analysis* |  |  |  |
| 24. Number of data coders | How many data coders coded the data? | Rapid qualitative methods were used, rather than traditional coding methodology. Two analysts reviewed the interview summary for each interview, then the primary analyst transferred findings into an Excel matrix for analysis. | 7 |
| 25. Description of coding tree | Did authors provide a description of the coding tree? | N/A, see above | 7 |
| 26. Derivation of themes | Were themes identified in advance or derived from the data? | Thematic analysis was not conducted. Domains regarding visual design were derived a posteriori. | 7 |
| 27. Software | Were themes identified in advance or derived from the data? | Microsoft Office Word and Excel (Version 2106, Build 14131.20332) | 7 |
| 28. Participant checking | Did participants provide feedback on the findings? | Participants did not provide feedback. | - |
| *Reporting* |  |  |  |
| 29. Quotations presented | Were participant quotations presented to illustrate the themes/findings?<br>Was each quotation identified? e.g. | Yes, in the results | 9-14 |
|  | participant<br>number |  |  |
| 30. Data and findings<br>consistent | Was there<br>consistency<br>between the data<br>presented and the<br>findings? | Yes | 9-14 |
| 31. Clarity of major<br>themes | Were major<br>themes clearly<br>presented in the<br>findings? | Yes | 9-14 |
| 32. Clarity of minor<br>themes | Were minor<br>themes clearly<br>presented in the<br>findings? | No | - |

### Supplemental Item 2. Interview guide (subset of questions most relevant to this paper in *italics*)

1. *What is your professional role?*
2. I have some basic demographic questions that I’d like to ask. Would that be okay? Please feel free to decline to answer if you do not wish to do so.

a. *What is your gender identity?*
b. *What races or ethnicities do you identify with?*
3. What are the main areas of clinical uncertainty you face when treating trauma patients?
4. Are there resources available to address those areas of uncertainty?

a. [if yes] Which resources do you use? Why?
b. [if no] What kinds of resources would you like? Why?
5. *What do you think about trauma guidance?*
6. What are your thoughts about using trauma guidance in your specific practice setting?
7. *How can we improve the design and usability of trauma clinical guidance?*
8. *Here is an example of a current trauma clinical guideline (asked to reflect on Figure 1, then repeated for Figure 2)*.

a. *What do you like about this?*
b. *What don’t you like about this?*
9. *Imagine a world where you could design trauma guidance. Now describe what you envision*.
10. What else would you like to share with me about improving resources/guidance for emergency medicine providers caring for trauma patients?

